# Novel Genetic Variants of Thoracic Aortic Aneurysm and Dissection: Evidence from a Japanese Community-Based Cohort

**DOI:** 10.1101/2025.09.17.25336030

**Authors:** Kentaro Akabane, Ken Nakamura, Hidenori Sato, Tsuneo Konta, Shusuke Arai, Yasushi Imai, Tetsuro Uchida

**Affiliations:** Second Department of Surgery, Yamagata University Faculty of Medicine, Yamagata, Japan; Division of Cardiovascular Surgery, Nihonkai General Hospital, Sakata, Japan; Department of Multiomics Research, Well-Being Research Institute, Yamagata University Faculty of Medicine, Yamagata, Japan; Department of Public Health and Hygiene, Yamagata University Faculty of Medicine, Yamagata, Japan; Division of Clinical Pharmacology, Department of Pharmacology, Jichi Medical University, Tochigi, Japan

**Keywords:** thoracic aortic aneurysm and dissection, novel genetic variant, Community-based Cohort, risk stratification, preventive strategies

## Abstract

**Background:** Thoracic aortic aneurysm and dissection (TAAD) is a life-threatening condition for which early risk stratification and preventive strategies represent critical challenges in modern medicine. Although genetic contributions have been well-established in high-risk populations, the clinical relevance of rare variants in the general population remains poorly understood. This study aimed to investigate their clinical significance using a community-based cohort.

**Methods:** We conducted a population-based survival analysis using the Yamagata Cohort, a prospective study in Japan. We selected 14 single-nucleotide polymorphisms from genes with definitive or strong clinical validity for TAAD, based on the criterion that the frequency of individuals homozygous for the minor allele was < 5%. Participants were categorized as carriers if they harbored homozygous rare variants, and as non-carriers otherwise. The primary outcome was TAAD-related mortality.

**Results:** Among 24,478 participants, we analyzed 5,722 individuals with genome-wide genotyping data. The carrier group included 1,499 individuals, and the non-carrier group comprised 4,223 individuals. TAAD-related deaths occurred in 12 individuals (8 carriers vs 4 non-carriers). The carrier group showed significantly lower survival rates than the non-carrier group (*P*=0.0010). In the multivariable Cox model, carrier status was independently associated with increased TAAD-related mortality (hazard ratio, 2.27; 95% confidence interval, 1.24–4.14; *P*=0.0056).

**Conclusions:** Rare homozygous variants in specific TAAD-related genes were significantly associated with TAAD-related mortality in this general Japanese population, despite the absence of prior pathogenic classification. These findings provide novel insights for pre-symptomatic risk stratification and a foundation for developing future preventive strategies in TAAD.

**Research Perspective:** *What New Question Does This Study Raise?:* Whether previously unclassified rare homozygous variants in definitive or strong TAAD-related genes contribute to disease-related mortality in the general population.

*What Question Should be Addressed Next?:* Future studies should determine whether these unclassified variants have true pathogenic roles or represent linkage with causal alleles, through functional validation and replication in other cohorts.

*What Are the Broader Clinical Implications?:* These findings may provide a foundation for novel clinical evaluation in general population, ultimately contributing to risk stratification and preventive interventions that could shift management strategies for TAAD.

## Introduction

Thoracic aortic aneurysm and dissection (TAAD) is a life-threatening cardiovascular disorder. A thoracic aortic aneurysm (TAA) is characterized by the progressive weakening and abnormal dilation of the aortic wall, which may ultimately lead to rupture or dissection. In contrast, aortic dissection involves a tear in the aortic intima that allows blood to penetrate the medial layer, forming a false lumen and potentially causing malperfusion or rupture. The incidence of TAAs is estimated at 5–10 cases per 100,000 person-years and that of aortic dissection at 5–30 per million people annually, indicating that these conditions are relatively uncommon but not exceedingly rare.^1^ Nonetheless, their fatality rates are extremely high, particularly for patients with untreated acute type A aortic dissection, where mortality increases by 2.6% per hour during the initial 24 hours and reaches up to 55% within 48 hours.^2^

As many individuals remain asymptomatic until catastrophic events occur, early risk stratification and prevention strategies are essential. Conventional risk factors such as hypertension, aging, and smoking are well-known contributors; however, they do not fully explain the development of TAAD, especially among younger individuals or in those without typical cardiovascular risk profiles. Notably, a strong heritable component has been recognized. Recent population-based studies have shown that first-degree relatives of individuals with aortic aneurysm or dissection have approximately a 6-fold increased risk of developing aneurysm and a 9-fold increased risk of dissection.^3^ In addition, the heritability of TAAD is estimated to be approximately 20%.^4^

To date, over 30 TAAD-related genes have been identified, many of which involve abnormalities in genes that encode vascular smooth muscle contractile proteins (e.g., *ACTA2*, *MYH11*, *MYLK*) or dysregulation of the transforming growth factor-beta (TGF-β) signaling pathway (e.g., *FBN1*, *TGFBR1*, *TGFBR2*, *SMAD3*).^5^ Among these, at least 11 genes have definitive or strong clinical validity for heritable TAAD, and approximately 25% of patients with familial or early-onset TAAD carry pathogenic variants in these genes.^6^ Despite these discoveries, most studies have focused on high-risk or syndromic populations, and the clinical significance of rare variants in the general population remains largely unexplored. Recent studies in various complex diseases have reported that rare variants with low minor allele frequency (MAF) can contribute to the genetic architecture of disease. ^7–9^ Moreover, the majority of TAAD-related mutations are dominantly inherited, although some severe early-onset cases have been linked to recessive variants such as homozygous *EFEMP2* mutations,^10^ suggesting that additional pathogenic mechanisms may remain undetected.

To address these unresolved questions, we conducted a population-based survival analysis derived from a community-based cohort study in Yamagata, Japan. We aimed to investigate whether individuals in the general population who carry rare variants in genes associated with TAAD have an increased risk of TAAD-related death. By linking genotypic data to long-term mortality outcomes, we sought to elucidate the clinical relevance of previously uncharacterized rare variants and identify novel genomic risk factors that could help in developing future strategies for risk stratification and prevention in TAAD.

## Methods

### Study design and participants

This study used data from the Yamagata Cohort, a prospective, community-based cohort study conducted in Yamagata Prefecture, Japan. Participants were enrolled during annual municipal health checkups conducted between January 2004 and December 2015. Eligible participants were residents of the prefecture, aged 39–74 years, who provided written informed consent for cohort participation, including future genetic analyses. Baseline data, including questionnaire responses (medical history, medication use, and lifestyle factors) and biospecimens, were collected. Individuals who did not consent to genetic testing or had incomplete baseline data were excluded from the present analysis. Participants were followed from enrollment until death or December 31, 2024, whichever occurred first. The study protocol was approved by the institutional review board of Yamagata University (Approval No. 2024-007). In accordance with ethical guidelines for secondary use of existing data and biospecimens, no additional consent was required for the present analysis. All procedures adhered to the principles of the Declaration of Helsinki.

### Genotyping and selection of single-nucleotide polymorphisms (SNPs)

Genomic DNA was extracted from peripheral blood samples. Genotyping was performed using three microarrays—Illumina Human Omni ExpressExome array, Illumina 660W-Quad array (Illumina, San Diego, CA, USA), and Axiom Japonica array (Thermo Fisher Scientific, MA, USA)—following the manufacturer’s standard protocols (Illumina, San Diego, CA, USA). Only directly genotyped SNPs were included in the analysis; imputed variants were excluded owing to potential inaccuracies in imputation quality. Quality control was performed as previously described.^11^ Specifically, SNPs with call rates below 95% or deviations from Hardy–Weinberg equilibrium (*P*<1 × 10⁻⁶) were excluded.

After quality control, candidate SNPs were identified from genes with definitive or strong clinical validity for TAAD.^6^ A total of 98 SNPs in these genes were screened initially. As this study focused on rare variants, 14 SNPs were selected for the final analysis based on the criterion that the frequency of individuals homozygous for the minor allele was < 5% in the Yamagata Cohort (Figure 1).

**Figure 1.**
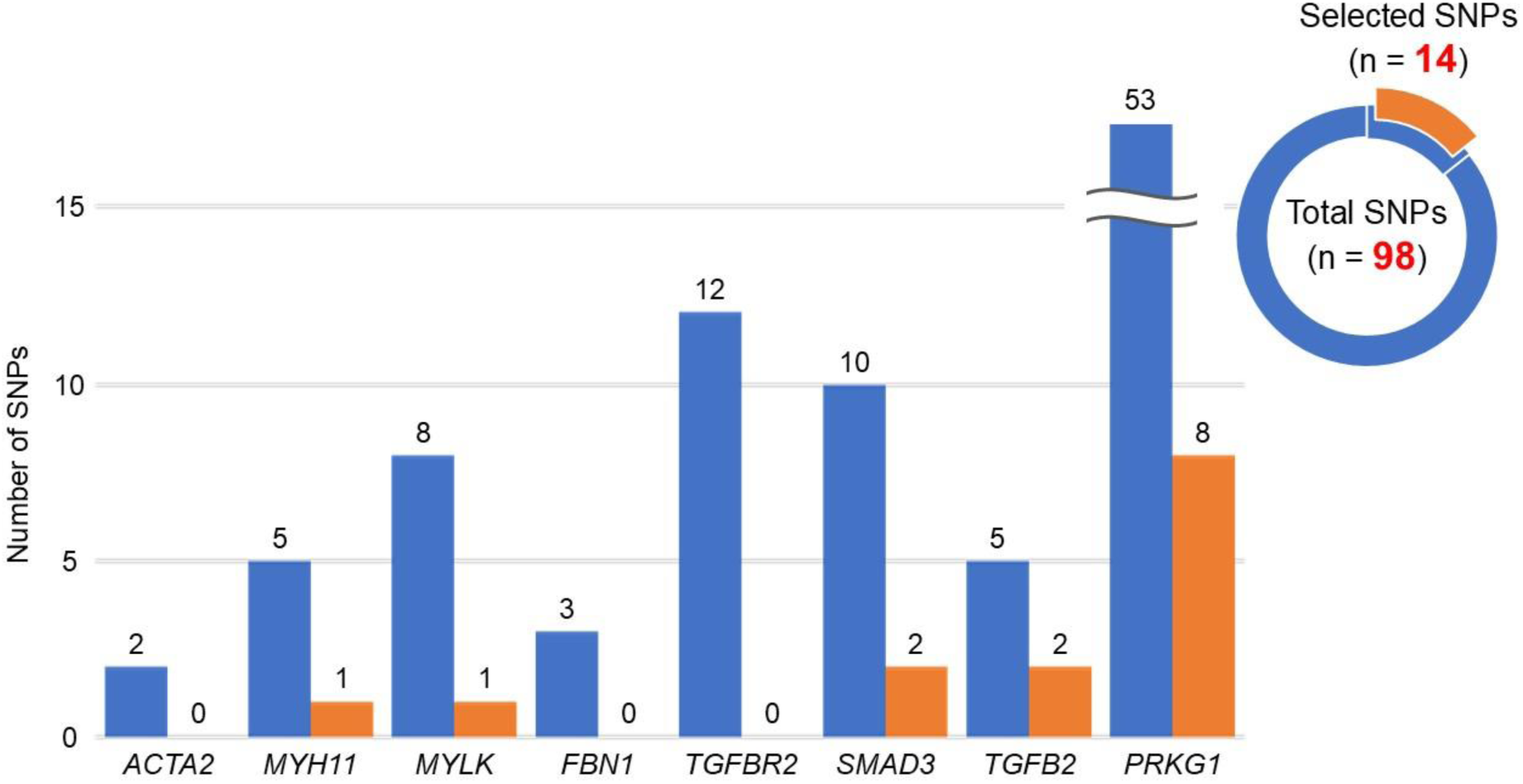
**Distribution of total and selected SNPs across TAAD-related genes.** A total of 98 SNPs in genes with definitive or strong clinical validity for TAAD were screened initially. Of these, 14 SNPs were selected for the final analysis based on the criterion that the frequency of individuals homozygous for the minor allele was < 5% in the Yamagata Cohort. SNP, single-nucleotide polymorphism; TAAD, thoracic aortic aneurysm and dissection

Functional predictions were obtained using CADD (v.1.7, GRCh38) and SpliceAI (v.1.3.1). For each variant, the CADD PHRED-scaled score and the maximum predicted Δ score from SpliceAI (across acceptor/donor gain/loss categories) were recorded. The ClinVar status was checked using the ClinVar database (accessed May 2025).

### Outcome definitions

The primary outcome was TAAD-related mortality, ascertained based on death certificate data obtained from the Vital Statistics Survey conducted by the Ministry of Health, Labour and Welfare of Japan. The causes of death were classified according to the International Classification of Diseases, 10th Revision (ICD-10). TAAD-related deaths were defined as those with ICD-10 code I71.0 (aortic dissection), I71.1 (ruptured thoracic aortic aneurysm), or I71.2 (unruptured thoracic aortic aneurysm).

### Definition of cardiovascular risk factors

Hypertension was defined as a systolic blood pressure (BP) ≥ 140 mmHg, diastolic BP ≥ 90 mmHg, or use of antihypertensive medications.^12^ Diabetes mellitus was defined as fasting plasma glucose (FPG) ≥ 126 mg/dL, hemoglobin A1c (HbA1c) ≥ 6.5%, or treatment with antidiabetic medication.^13^ Dyslipidemia was defined as low-density lipoprotein cholesterol (LDL-C) ≥ 140 mg/dL, high-density lipoprotein cholesterol (HDL-C) < 40 mg/dL, triglyceride (TG) ≥ 150 mg/dL, or use of lipid-lowering medications.^14^ Obesity was defined as a body mass index (BMI) of ≥ 25 kg/m².^15^

### Statistical analysis

Participants were assigned to the “carrier group” if they harbored a homozygous genotype for the minor allele in at least one of the 14 selected SNPs. The remaining participants were classified as the “non-carrier group.” Descriptive statistics are presented as means with standard deviations for continuous variables and as frequencies and percentages for categorical variables. Between-group comparisons were performed using the Mann–Whitney U test or the chi-square test, as appropriate. The Kaplan–Meier survival analysis was performed to compare TAAD-related mortality between the two groups, and differences were evaluated using the log-rank test. Multivariable Cox proportional hazards models were constructed to estimate hazard ratios (HRs) and 95% confidence intervals (CIs), adjusting for potential confounders including age, sex, smoking status, hypertension, diabetes, dyslipidemia, and obesity. All statistical analyses were performed using JMP v.18 (SAS Institute Inc., Cary, NC, USA). The corresponding author had full access to all the data in the study and takes responsibility for the integrity of the data and the accuracy of the analysis. The data that support the findings of this study are derived from the Yamagata Cohort. Due to institutional and ethical restrictions, these datasets are not publicly available. However, de-identified data may be shared by the corresponding author upon reasonable request and with approval of the institutional review board, in accordance with the AHA Journals’ implementation of the Transparency and Openness Promotion (TOP) Guidelines.

## Results

### Study population and variant characteristics

Of the 24,478 participants, 5,948 underwent genome-wide genotyping. After excluding those with missing baseline clinical data, 5,722 individuals were included in the final analysis. All participants were prospectively followed up for a mean period of 5,237±1,326 days. Vital status was available for all participants through linkage to the municipal death registry, ensuring complete follow-up. The carrier group included 1,499 participants (26.2%), and the non-carrier group comprised 4,223 participants (73.8%) (Figure 2). Significant differences between the two groups were observed with regard to age (64.0±9.4 vs. 63.3±9.2 years, *P*=0.0005), diastolic BP (76.8±10.5 vs. 77.7±10.9, *P*=0.0011), prevalence of dyslipidemia (725 [48.4%] vs. 2177 [51.6%], *P*=0.0341), and LDL-C levels (125.0±30.7 vs. 127.0±30.2, *P*=0.0497). Additional baseline characteristics are summarized in Table 1.

**Figure 2.**
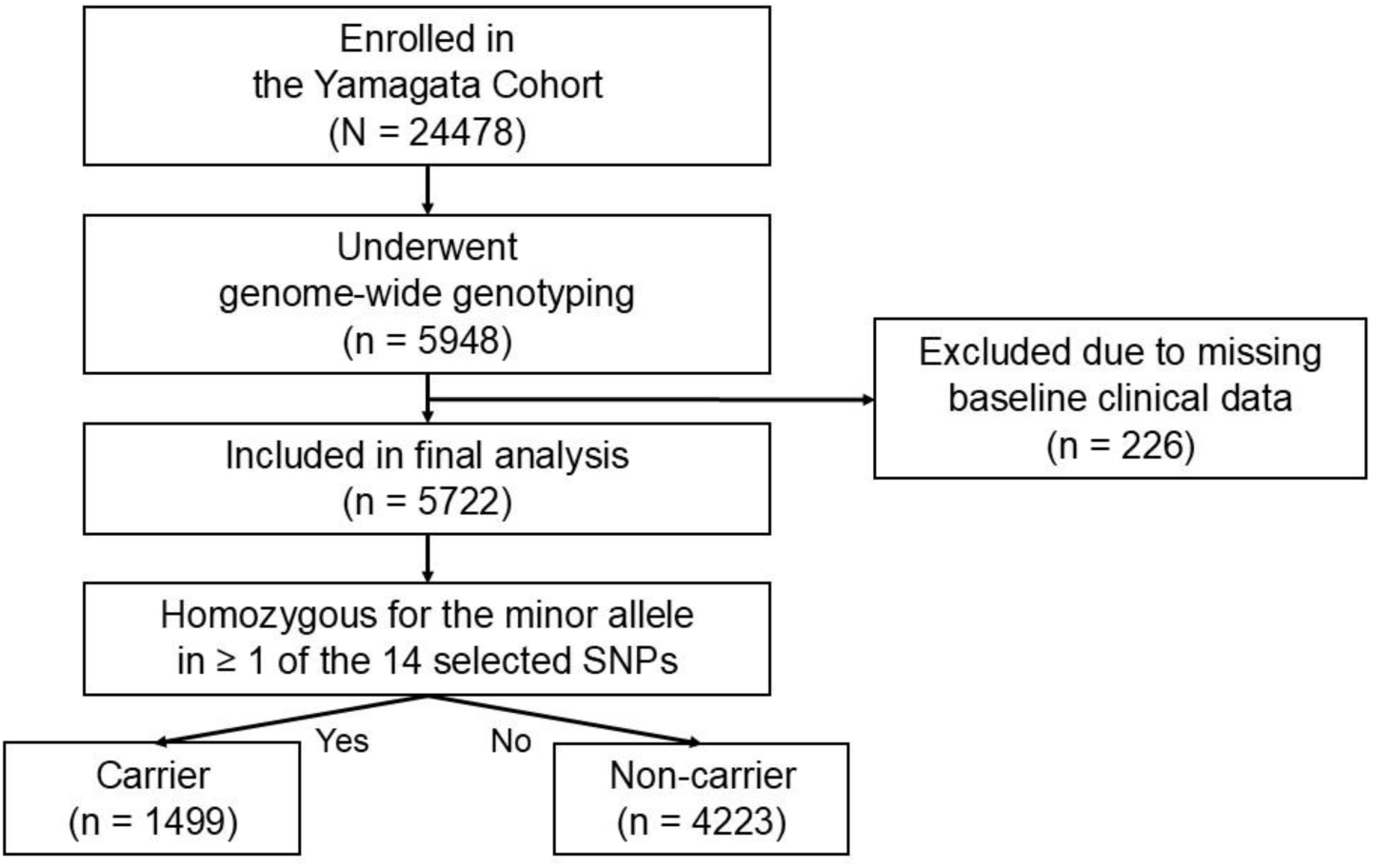
**Participant selection flowchart.** Of the 24,478 participants, 5,948 underwent genome-wide genotyping. After excluding those with missing baseline clinical data, 5,722 individuals were included in the final analysis. The carrier group included 1,499 participants (26.2%), and the non-carrier group comprised 4,223 participants (73.8%).

**Table 1.** Baseline characteristics of the study population.

|  | Carrier (n = 1499) | Non-carrier (n = 4223) | *P* value |
| --- | --- | --- | --- |
| Age, y, mean (SD) | 64.0 (9.4) | 63.3 (9.2) | 0.0005 |
| Male sex, n (%) | 681 (45.4) | 1946 (46.1) | 0.6640 |
| Smoking history, n (%) | 486 (32.4) | 1367 (32.4) | 0.9709 |
| Hypertension, n (%) | 642 (42.8) | 1845 (43.7) | 0.5634 |
| Systolic BP, mmHg, mean (SD) | 129.5 (17.2) | 130.2 (17.4) | 0.1067 |
| Diastolic BP, mmHg, mean (SD) | 76.8 (10.5) | 77.7 (10.9) | 0.0011 |
| Diabetes mellitus, n (%) | 301 (20.1) | 800 (18.9) | 0.3394 |
| FPG, mg/dL, mean (SD) | 97.2 (17.4) | 98.0 (20.4) | 0.4016 |
| HbA1c, %, mean (SD) | 5.3 (0.6) | 5.3 (0.6) | 0.4114 |
| Dyslipidemia, n (%) | 725 (48.4) | 2177 (51.6) | 0.0341 |
| LDL-C, mg/dL, mean (SD) | 125.0 (30.7) | 127.0 (30.2) | 0.0497 |
| HDL-C, mg/dL, mean (SD) | 61.6 (15.8) | 61.7 (15.3) | 0.8238 |
| TG, mg/dL, mean (SD) | 116.8 (75.0) | 117.3 (75.6) | 0.6563 |
| Obesity, n (%) | 417 (27.8) | 1170 (27.7) | 0.9330 |
| BMI, kg/m<sup>2</sup>, mean (SD) | 23.3 (3.2) | 23.3 (3.2) | 0.6468 |

Values are expressed as mean (standard deviation) for continuous variables and number (%) for categorical variables. *P* values were calculated using the Mann–Whitney U test for continuous variables and the chi-square test for categorical variables.
BMI, body mass index; BP, blood pressure; FPG, fasting plasma glucose; HbA1c, glycosylated hemoglobin A1c; HDL-C, high-density lipoprotein cholesterol; LDL-C, low-density lipoprotein cholesterol; SD, standard deviation; TG, triglycerides.

A variant was identified at rs11647019 (T > C) in MYH11 and at rs820363 (A > G) in MYLK, both of which encode vascular smooth muscle contractile proteins. In addition, a total of eight rare homozygous variants were detected in PRKG1, another gene involved in vascular smooth muscle contraction, including rs10508937 (C > T), rs10490969 (T > C), rs7091469 (C > T), rs12766318 (G > A), rs10997716 (A > G), rs293312 (C > T), rs10762176 (C > T), and rs2001283 (G > A). Variants were also identified in *SMAD3* (rs920293 [A > T], rs2118610 [C > T]) and *TGFB2* (rs10482750 [T > C], rs12058490 [A > G]), genes that encode components of the TGF-β signaling pathway. All 14 SNPs selected for analyses were unregistered in ClinVar and showed low predicted pathogenicity based on CADD (PHRED-scaled scores < 15) and SpliceAI (maximum Δ scores < 0.1). The list of identified SNPs is presented in Table 2.

**Table 2.**
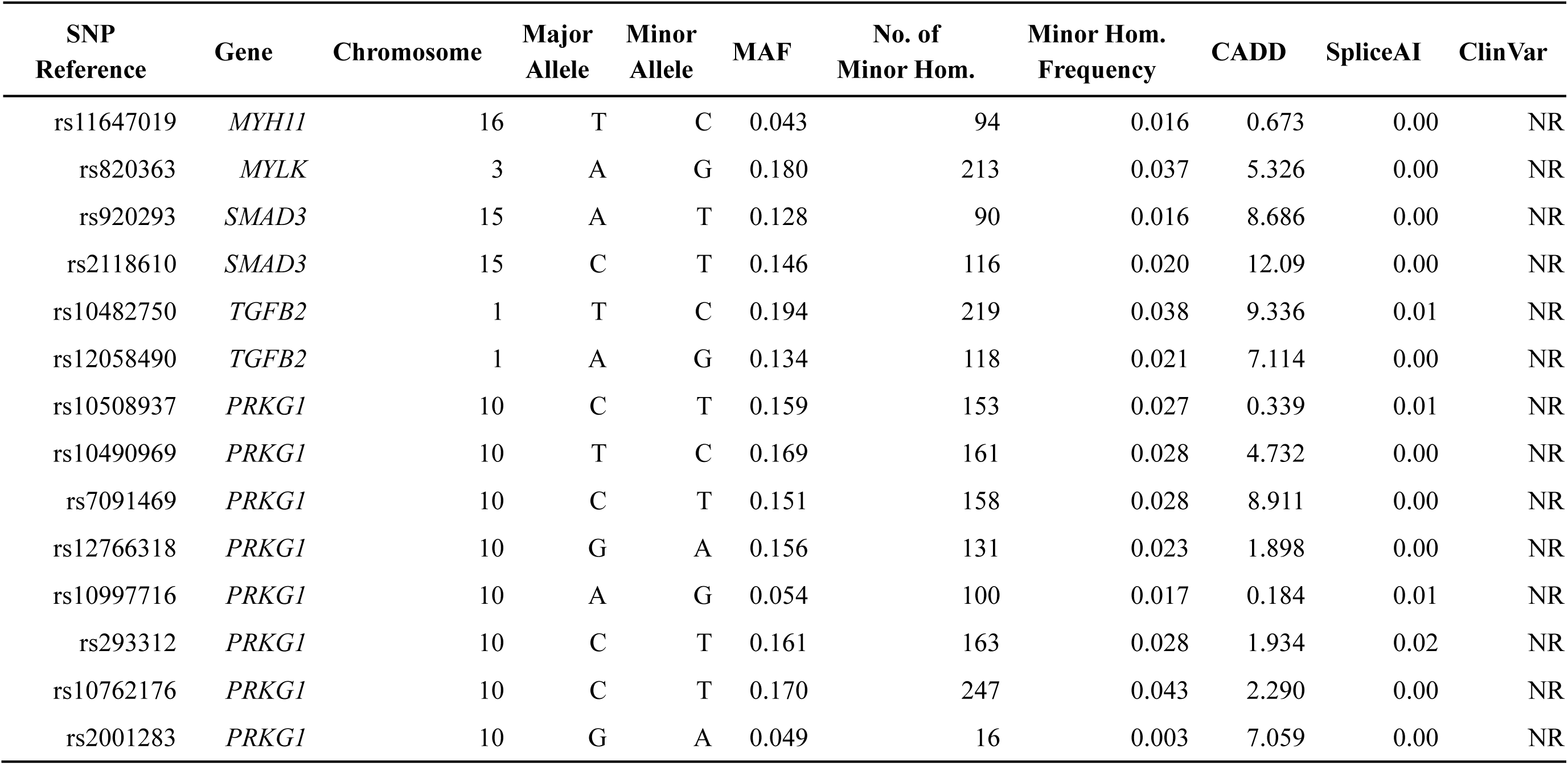
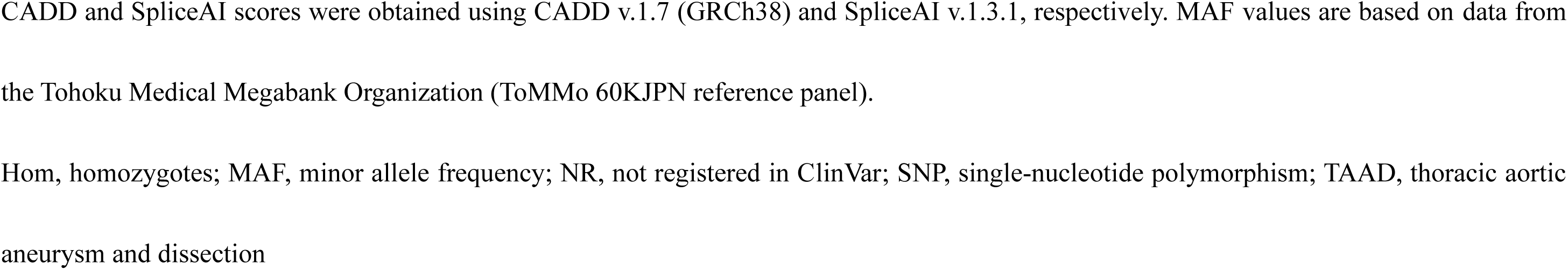
Summary of 14 rare homozygous SNPs in TAAD-related genes.

### Kaplan–Meier survival analysis

TAAD-related deaths occurred in 12 individuals: 8 (0.53%) in the carrier group and 4 (0.09%) in the non-carrier group. Kaplan–Meier survival curves showed a significantly lower cumulative survival in the carrier group than in the non-carrier group. The log-rank test revealed a significant difference in TAAD-related mortality between the two groups (*P*=0.0010). Survival curves diverged clearly after approximately 4,000 days of follow-up (Figure 3).

**Figure 3.**
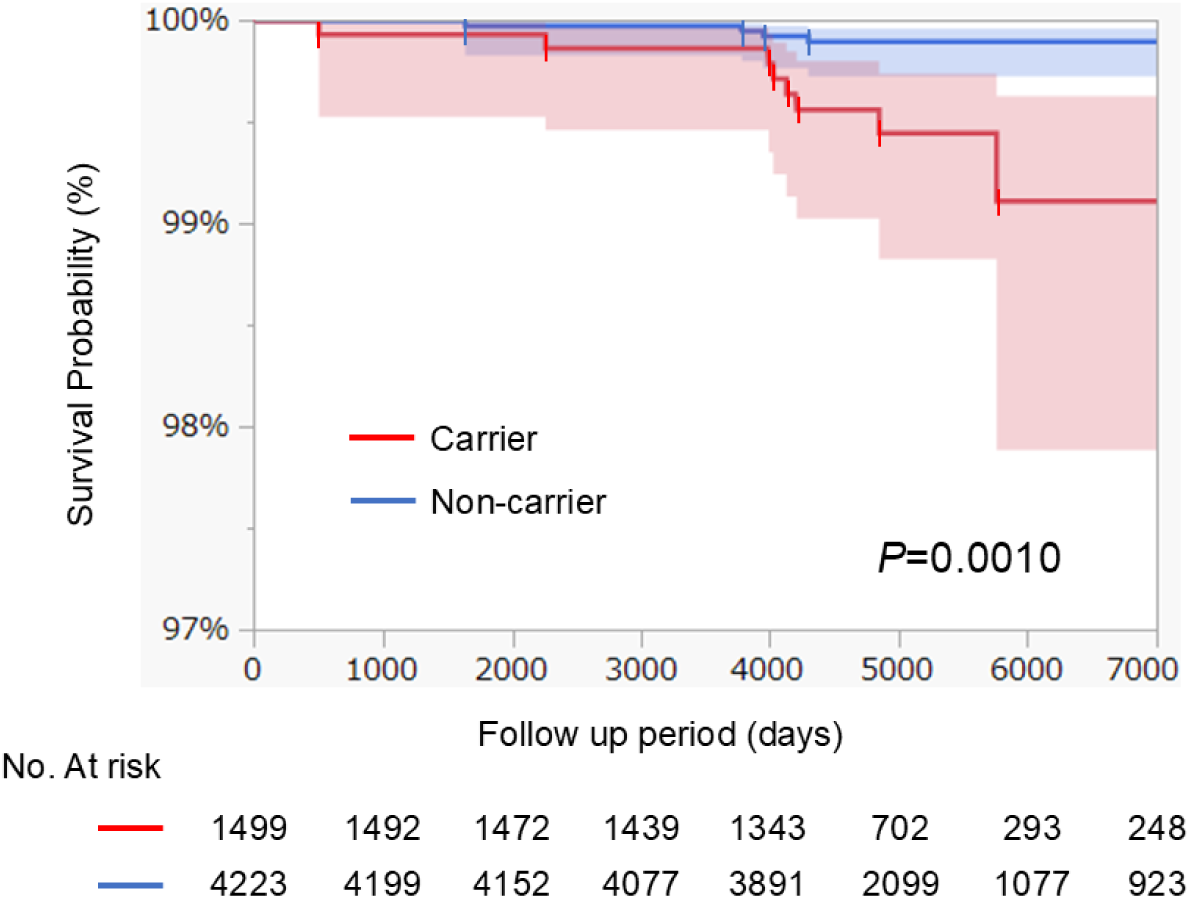
**TAAD-related mortality according to SNP carrier status.** Kaplan–Meier survival curves showed a significantly lower cumulative survival in the carrier group than in the non-carrier group. The log-rank test revealed a significant difference in TAAD-related mortality between the two groups (*P*=0.0010). SNP, single-nucleotide polymorphism; TAAD, thoracic aortic aneurysm and dissection

### Cox proportional hazards analysis

In a multivariable Cox proportional hazards model adjusting for age, sex, smoking history, hypertension, diabetes mellitus, dyslipidemia, and obesity, the carrier status remained significantly associated with TAAD-related mortality (HR, 2.27; 95% CI, 1.24–4.14; *P*=0.0056). Notably, age (HR, 1.15; 95% CI, 1.04–1.28; *P*=0.0015) and smoking history (HR, 2.34; 95% CI, 1.02–5.36; *P*=0.0385) were also significantly associated with increased risk, whereas hypertension (HR, 1.69; 95% CI, 0.87–3.31; *P*=0.1013), despite being a well-known risk factor, did not exhibit a significant association in this model (Figure 4).

**Figure 4.**
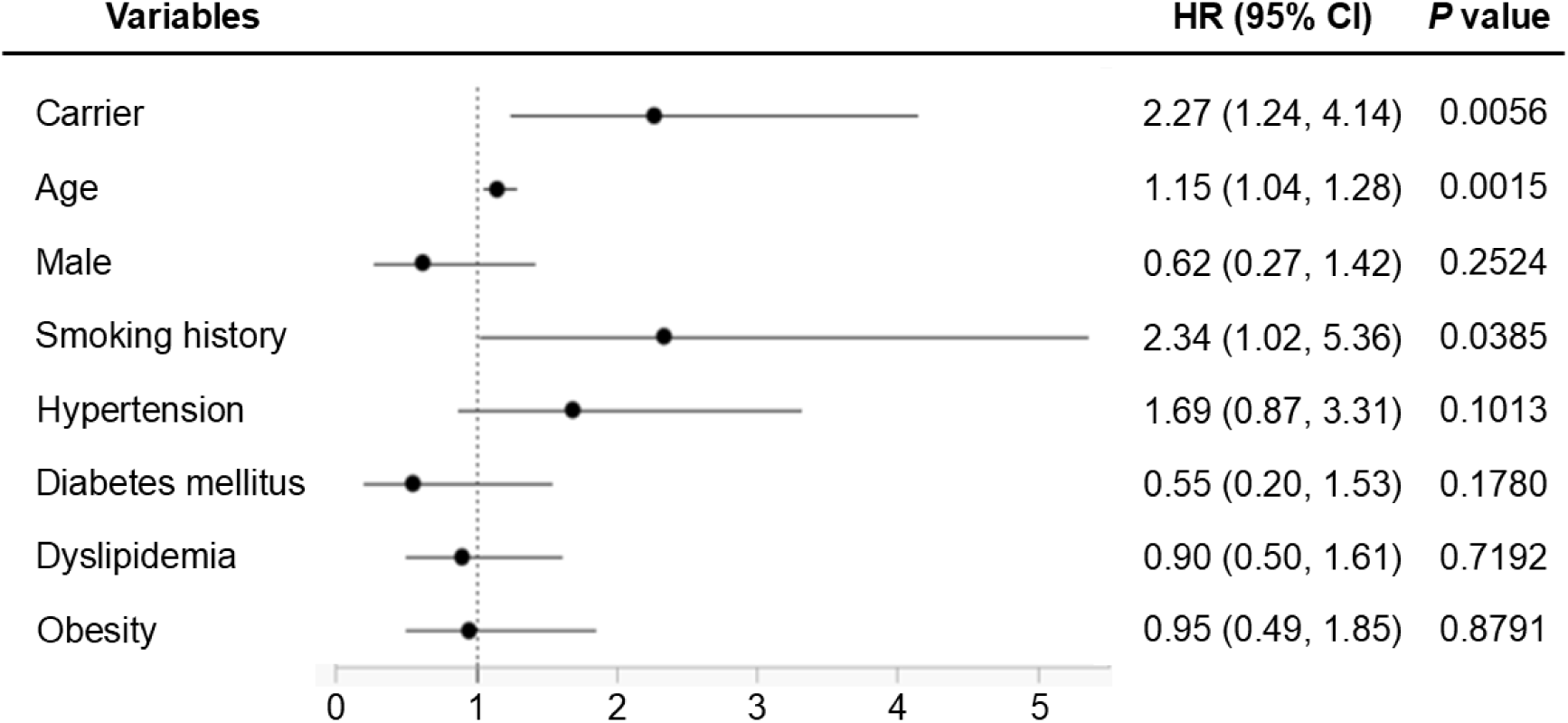
**Multivariable Cox proportional hazards model for TAAD-related mortality.** In a multivariable Cox proportional hazards model adjusted for age, sex, smoking history, hypertension, diabetes mellitus, dyslipidemia, and obesity, the carrier status remained significantly associated with TAAD-related mortality (HR, 2.27; 95% CI, 1.24–4.14; *P*=0.0056). SNP, single-nucleotide polymorphism; TAAD, thoracic aortic aneurysm and dissection; HR, hazard ratio; CI, confidence interval

## Discussion

This study showed that rare homozygous variants in specific TAAD-related genes were significantly associated with TAAD-related mortality in a large, community-based Japanese cohort. Notably, none of the analyzed SNPs had been registered in ClinVar, and all exhibited low predicted pathogenicity based on in silico tools. Nonetheless, carriers exhibited a markedly higher risk of death than non-carriers. These findings are consistent with those from recent reports suggesting that, beyond known pathogenic variants, some rare variants that are currently unclassified may also increase disease risk^16–18^; this implies that clinically relevant signals may be embedded within these unclassified variants.

Most genetic studies on TAAD have focused primarily on high-risk populations, such as patients with syndromic features (e.g., Marfan or Loeys–Dietz syndrome), patients with early-onset disease, or those with a positive family history. As a result, our understanding of the genetic basis of TAAD has been largely shaped by pathogenic variants identified in these high-risk populations.^19,20^ These studies have contributed substantially to identifying causative genes; however, they do not fully represent the genetic landscape of sporadic or asymptomatic individuals in the general population. Indeed, despite approximately 80% of TAAD cases being considered sporadic, genetic testing is not routinely performed in patients without syndromic features or a suggestive family history.^20,21^ Although some evidence suggests that even non-syndromic cases may have pathogenic or risk-associated variants,^22^ the clinical significance of such variants remains poorly understood because there are few large-scale population-based studies that integrate genomic profiles and longitudinal outcomes.

In this context, the present study uniquely focused on a community-based cohort without prior clinical selection criteria, enabling an unbiased investigation of genetic risk in the general population. By integrating genotypic data with long-term survival outcomes, our analysis offers novel insights into the relevance of rare variants that are currently unclassified in public databases, particularly among individuals who would not typically undergo genetic screening. This approach presents a practical alternative to traditional methods such as cosegregation analysis and experimental validation, which are often unfeasible for low-frequency variants in population-based settings. By leveraging survival analysis, we detected significant associations even in the absence of prior functional data, highlighting a potential pathway for prioritizing candidate variants for further study.

Although none of the identified SNPs in this study had been previously classified as pathogenic, their significant association with TAAD-related mortality suggests that certain uncharacterized variants may either pose a pathogenic risk themselves or be in linkage disequilibrium with true causal alleles. This analysis is especially valuable in situations where there is a lack of functional or experimental data and could help improve the classification of unclassified variants in TAAD.

In summary, our findings highlight the utility of integrating genomic and phenotypic data in population-based studies to identify novel genetic risk factors. These results lay a foundation for future research to validate the observed associations, clarify underlying biological mechanisms, and integrate these variants into risk stratification and preventive strategies for TAAD.

## Limitations

This study has several limitations that should be acknowledged. First, the number of TAAD-related deaths was extremely small (n = 12), which limits statistical power and warrants cautious interpretation of the associations observed. In addition, causes of death were determined solely from ICD-10 codes on death certificates, and detailed clinical information on TAAD diagnosis, treatment history, or symptom onset was unavailable for deceased individuals. Second, although 98 SNPs in definitive or strong TAAD-related genes were initially screened, only 14 rare homozygous variants were included in the final analysis. As a result, established TAAD-related genes such as *ACTA2* were excluded owing to a lack of qualifying variants, potentially introducing selection bias and limiting generalizability. Moreover, because the analysis was based on a combined set of qualifying SNPs across multiple genes, this study design did not allow for determination of the relative contribution of each individual variant to TAAD-related mortality. Further variant-level analyses are needed to clarify their respective roles. Third, functional validation of the selected SNPs at the experimental level was not conducted. Although in silico tools such as CADD and SpliceAI were used for annotation, they alone cannot establish pathogenicity or exclude linkage disequilibrium with causal alleles. Finally, this was a single-center, population-based study conducted in Japan, and the findings require external validation in other cohorts to confirm their broader applicability.

## Conclusions

This study identified a significant association between rare homozygous variants in definitive or strong TAAD-related genes and TAAD-related mortality in a large, community-based Japanese cohort. Although none of the analyzed SNPs had been previously reported as pathogenic, carriers demonstrated a significantly higher risk of TAAD-related deaths. Our findings suggest that some unclassified variants may have unrecognized pathogenic relevance. By integrating genomic screening with long-term outcome data, our method provides a practical strategy for uncovering previously unrecognized genetic risk factors. This may contribute to future efforts in early risk stratification, and preventive intervention for patients with TAAD.

## Data Availability

The data underlying this study are derived from the Yamagata Cohort and cannot be made publicly available due to ethical and legal restrictions. Summary-level results are provided within the article and its supplementary material. Additional data may be available from the corresponding author upon reasonable request and subject to institutional ethics committee approval.

## Acknowledgments

The authors would like to thank the members of the Second Department of Surgery for their valuable advice and support in the preparation of this manuscript.

## Sources of Funding

This research received no specific grant from any funding agency.

## Disclosures

The authors have no conflicts of interest to disclose.

## Non-standard Abbreviations and Acronyms

BMI: Body Mass Index
BP: Blood Pressure
CADD: Combined Annotation Dependent Depletion
CI: Confidence Interval
ClinVar: Clinical Variant Database
FPG: Fasting Plasma Glucose
HbA1c: Hemoglobin A1c
HDL-C: High-Density Lipoprotein Cholesterol
HR: Hazard Ratio
LDL-C: Low-Density Lipoprotein Cholesterol
MAF: Minor Allele Frequency
NR: Not Registered (in ClinVar)
SNP: Single-Nucleotide Polymorphism
SpliceAI: Splicing Artificial Intelligence
TAAD: Thoracic Aortic Aneurysm and Dissection
TAA: Thoracic Aortic Aneurysm
TG: Triglyceride
YUGCC: Yamagata University Genomic Cohort Consortium

## Notes

### Competing Interest Statement

The authors have declared no competing interest.

### Clinical Trial

This study is a prospective observational cohort study and was not registered as a clinical trial because it does not involve any interventional component.

### Funding Statement

This research received no external funding.

### Author Declarations

The study protocol was approved by the institutional review board of Yamagata University (Approval No. 2024-007).

